# Mental health, personality and lifetime psychedelic use during the COVID-19 pandemic

**DOI:** 10.1101/2020.12.24.20248519

**Authors:** Federico Cavanna, Carla Pallavicini, Virginia Milano, Juan Cuiule, Rocco Di Tella, Pablo González, Enzo Tagliazucchi

**Affiliations:** Departamento de Física, Universidad de Buenos Aires and Instituto de Física de Buenos Aires (IFIBA – CONICET), Pabellón I, Ciudad Universitaria (1428), CABA, Buenos Aires, Argentina; Fundación para la lucha contra las enfermedades neurológicas de la infancia (FLENI), Montañeses 2325, C1428 CABA, Buenos Aires, Argentina; El Gato y La Caja, Buenos Aires, Argentina, Teodoro García 2747, C1426DMU CABA

**Keywords:** psychedelics, mental health, resilience, well-being, personality traits, COVID-19

## Abstract

**Background:** The COVID-19 pandemic and its consequences represent a major challenge to the mental health and well-being of the general population. Some groups may be more vulnerable than others, depending on factors such as preexisting conditions, personality, and past life experiences. Building on previous work on the potential long-term benefits of psychedelics, we hypothesized that lifetime use of these drugs could be linked to better mental health indicators in the context of the ongoing pandemic.

**Methods:** Two anonymous online surveys were conducted between April 2020 and June 2020, including questions about lifetime experience with psychedelics and other psychoactive drugs, and psychometric scales designed to measure personality traits, anxiety, negative and positive affect, well-being and resilience. Principal component analysis was applied to divide the sample into groups of subjects based on their drug use reports.

**Results:** 5618 participants (29.15 ± 0.12 years, 71.97% female) completed both surveys and met the inclusion criteria, with 32.43% of the final sample reporting at least one use of a psychedelic drug. Lifetime psychedelic use was linked to increased openness and decreased conscientiousness, and with higher scores of positive affect. The reported number of past psychedelic experiences predicted higher scores of the secondary personality trait beta factor, which has been interpreted as a measure of plasticity. No significant associations between lifetime use of psychedelics and indicators of impaired mental health were observed.

**Conclusion:** We did not find evidence of an association between lifetime use of psychedelics and poor mental health indicators. Conversely, experience with psychedelic drugs was linked to increased positive affect and to personality traits that favor resilience and stability in the light of the ongoing crisis. Future studies should be conducted to investigate these results from a causal perspective.

## Introduction

Psychedelic drugs are found in nature in multiple species of plants, fungi and animals (Rätsch, 2005). The use of certain compounds (i.e. mescaline, psilocybin, N,N-dimethyltryptamine [DMT]) has been documented at least for centuries, mainly in association with medicinal, religious, and other ceremonial practices (e.g. divination) (Escohotado, 2002). The molecular structure of psychedelics resembles that of serotonin (an endogenous neurotransmitter), which enables their pharmacological action as 2A serotonin receptors (5-HT_2A_) agonists (Nichols, 2016). At the phenomenological, cognitive and behavioral levels, 5-HT_2A_ agonism is linked to a wide range of effects that alter the conscious experience of the user, such as modifications in the perception of the environment and the self, sedation, stimulation, as well as changes in mood, prosocial behavior, cognitive flexibility and creativity, among others (Schmid et al., 2015; Nichols, 2016; Preller and Vollwenweider, 2016; Jungaberle et al., 2018; Palamar and Acosta, 2020). These effects attracted the attention of mainstream science during the 1950s, leading to several investigations in healthy and clinical populations (Dyck and Farrell, 2018). However, the widespread availability of psychedelic drugs (mainly LSD) during the 1950s and 1960s, combined with a complex social and political landscape, led to the classification of several psychedelics as Schedule 1 drugs, which effectively shut down most research on these substances and their potential clinical uses (Rucker et al., 2018).

The prohibition on psychedelic drugs was partially justified by concerns regarding potential long-term negative effects on mental health (Lee and Shlain, 1992). However, subsequent studies failed to establish a link between lifetime use of psychedelic drugs and increased rate of mental health issues. Krebs and Johansen analyzed data from 130.152 randomly selected individuals, of which 21.967 reported at least one experience with psychedelic drugs (Krebs and Johansen, 2013). Their study failed to detect significant associations between psychedelic use and several markers of impaired mental health, including serious psychological distress, inpatient or outpatient mental health treatment, and symptoms of nine psychiatric disorders. A follow-up study by the same authors found the same results after adjusting for sociodemographics, the use of other drugs, and childhood depression (Johansen and Krebs, 2015). Converging results have been published for controlled laboratory studies (Nichols, 2016). Assessments by interdisciplinary panels of experts consistently ranked psychedelics as some of the least harmful recreational drugs, with safety profiles substantially better than those of widely available and consumed drugs, such as alcohol and tobacco (Nutt et al., 2010).

In recent years, the scientific and clinical interest in psychedelics has increased considerably as a consequence of a new wave of research demonstrating their potential in neuroscience, psychiatry, and as adjuncts for psychotherapy (Kleber, 2016; Johnson and Griffiths, 2017; Nichols and Hendricks, 2020; Nutt and Carhart-Harris, 2020; Reiff et al., 2020). One of the major promises of psychedelics as therapeutic agents is their capacity to induce long-term psychological and behavioral changes after a single session (MacLean et al., 2011; Lebedev et al., 2016; Bouso et al., 2018; Erritzoe et al., 2018; Erritzoe et al., 2019), which highlights their enormous potential in the treatment of certain mood disorders and addictive behaviors (Bogenschutz et al., 2015; Johnson et al., 2014; Johnson et al., 2017). Crucially, these effects appear to be mediated by the nature of the induced psychedelic experience, with reports of mystical-type experiences being associated with better outcomes (Garcia-Romeu et al., 2014; Griffiths et al., 2018; Johnson et al., 2019). While these effects have been shown for specific clinical populations, it could be hypothesized that psychedelic use might lead to long-term mental health benefits in healthy individuals. These benefits might remain unnoticed until the individual is challenged by adverse personal or social circumstances.

We investigated the relationship between lifetime psychedelic use and multiple mental health indicators in the context of the COVID-19 pandemic, an adverse event of worldwide scope. COVID-19 is a contagious respiratory and vascular disease caused by acute respiratory syndrome coronavirus 2 (SARS-CoV-2). The rapid spread of COVID-19 during early 2020 precipitated drastic lockdown measures in several countries, which in turn impacted negatively in the mental health of the population (Pfefferbaum and North, 2020); in particular, this study was conducted in Argentina, where one of the longest lockdowns was declared between March and November. Our first main objective was to determine differences in mental health indicators (anxiety, positive and negative affect, well-being, resilience) between individuals who reported past psychedelic use, and those who declared their past use of other psychoactive drugs. Our second objective was to investigate the potential impact of lifetime psychedelic use on personality traits, and how this potential impact related to different mental health indicators.

## Materials and methods

### Survey

An anonymous Internet-based survey with two parts was conducted between April 2020 and June 2020, i.e. between one and three months after a severe lockdown was declared in most of Argentina’s provinces. Recruitment advertisements were shared via social media web pages (e.g. Facebook, Instagram and Twitter). The first part of the survey was promoted as a questionnaire to understand the relationship between psychoactive drug use and mental health during the COVID-19 pandemic, while the second part was promoted as a questionnaire to determine personality traits. The survey was presented in Spanish and each of its parts took approximately 20 minutes to complete. All subjects provided informed consent and received no compensation for their participation. The procedure was included within a larger protocol for online experimentation approved by the ethics committee of Centro de Educación Médica e Investigaciones Clínicas Norberto Quirno.

### Questionnaires and scales

The full survey is available online (https://investigacion.elgatoylacaja.com/concienciaysustancia/ and https://investigacion.elgatoylacaja.com/personalidad/). The first questionnaire collected sociodemographic variables (age, gender and nationality), variables related to past use of psychoactive compounds (drugs that were consumed by the user and their frequency of use), and the STAI, PANAS, BIEPS and RS scales. The second part of the survey assessed personality via the BFI questionnaire.

State-Trait Anxiety Inventory (STAI): a commonly used scale which measures state anxiety (which refers to a situational anxiety of a temporary nature) and trait anxiety (which designates a stable trait linked to personal characteristics) (Spielberger et al., 1983). The instrument comprises 40 items and is based on a 4-point Likert scale, ranging from “Almost Never” to “Almost Always’’.

Positive and Negative Affect Schedule (PANAS): a psychometric scale that has been widely used to measure both proposed dimensions of affect, positive and negative (Watson et al., 1988). The instrument consists of 20 affirmations based on a 5-point Likert scale, ranging from “Not at all” to “Very much”.

Psychological Well-being Scale (BIEPS): a scale used to measure eudaimonic well-being in adults (including dimensions of acceptance, perception of control, social ties, and autonomy and projects) (Casullo & Brenlla, 2002). It consists of 13 questions based on a 3-point Likert scale, ranging from “Disagree” to “Agree”.

Resilience Scale (RS): a questionnaire aiming to evaluate Resilience through the subscales of self-reliance, purpose capacity and life meaning and cognitive avoidance (Wagnild & Young, 1993). In this study a local adaptation was used (Rodríguez et al., 2009) which consists of 21 items based on a 5-point Likert scale, ranging from “Fully agree” to “Fully disagree”.

Big Five Inventory (BFI): an inventory assessing five dimensions of personality: neuroticism, extraversion, openness to experience, agreeableness, and conscientiousness (Benet-Martinez and John, 1998). The BFI questionnaire consists of 44 items based on a 5-point Likert scale, ranging from “Fully disagree” to “Fully agree”.

### Data processing and analysis

Data from the two questionnaires was merged according to a unique identifier. Due to the scope of the ethics approval, and because the local adaptation of psychometric scales is limited to male and female Argentinian adults, only participants older than 18 years old, residents of Argentina, and with male or female gender identity were retained for subsequent analysis.

Psychometric scales (i.e., STAI, PANAS, BIEPS, RS, BFI) were scored according to gender and age, obtaining a statistical score adjusted to the local population. Next, all values were converted to a uniform scale between 0 and 10 in order to facilitate comparison.

Descriptive statistics (e.g. means, standard deviations and standard errors from the mean [SEM]) were used to characterize the scale and subscale scores of the survey. To reduce the dimensionality of the data and thus the number of independent statistical tests, we first conducted a principal component analysis (PCA) of the data based on computing the singular value decomposition of the centered data matrix. After grouping the drug use variables into interpretable principal components, we applied analysis of variance (ANOVA) followed by Student t-tests to determine statistically significant differences between groups of users assigned to the different principal components. Bonferroni correction for multiple comparisons was applied whenever stated in the description of the results.

## Results

During the data collection window (April 2020 - May 2020), 11.365 individuals answered the first part of the survey (demographics, drug use questionnaire, STAI, PANAS, BIEPS, RS) and 157.101 answered the second part (BFI questionnaire), with 10.722 participants answering both parts of the survey. Of these, 5.104 were excluded as they failed to meet the inclusion criteria presented in the “Materials and methods” section. Thus, the final sample consisted of 5.618 participants who were 29.15 (± 0.12) years old and 71.97% female. 32.43% of the total sample reported at least one use of a psychedelic drug.

The demographic information is summarized in Table 1. Demographic information per group of participants associated with each principal component (see below) is reported in Table 2 of the supplementary material. Note that percentages in the first row of Table 1 do not add to 100%, since reported uses of different drugs are not exclusive (e.g. one participant could have reported past uses of different drugs).

**Table 1.**
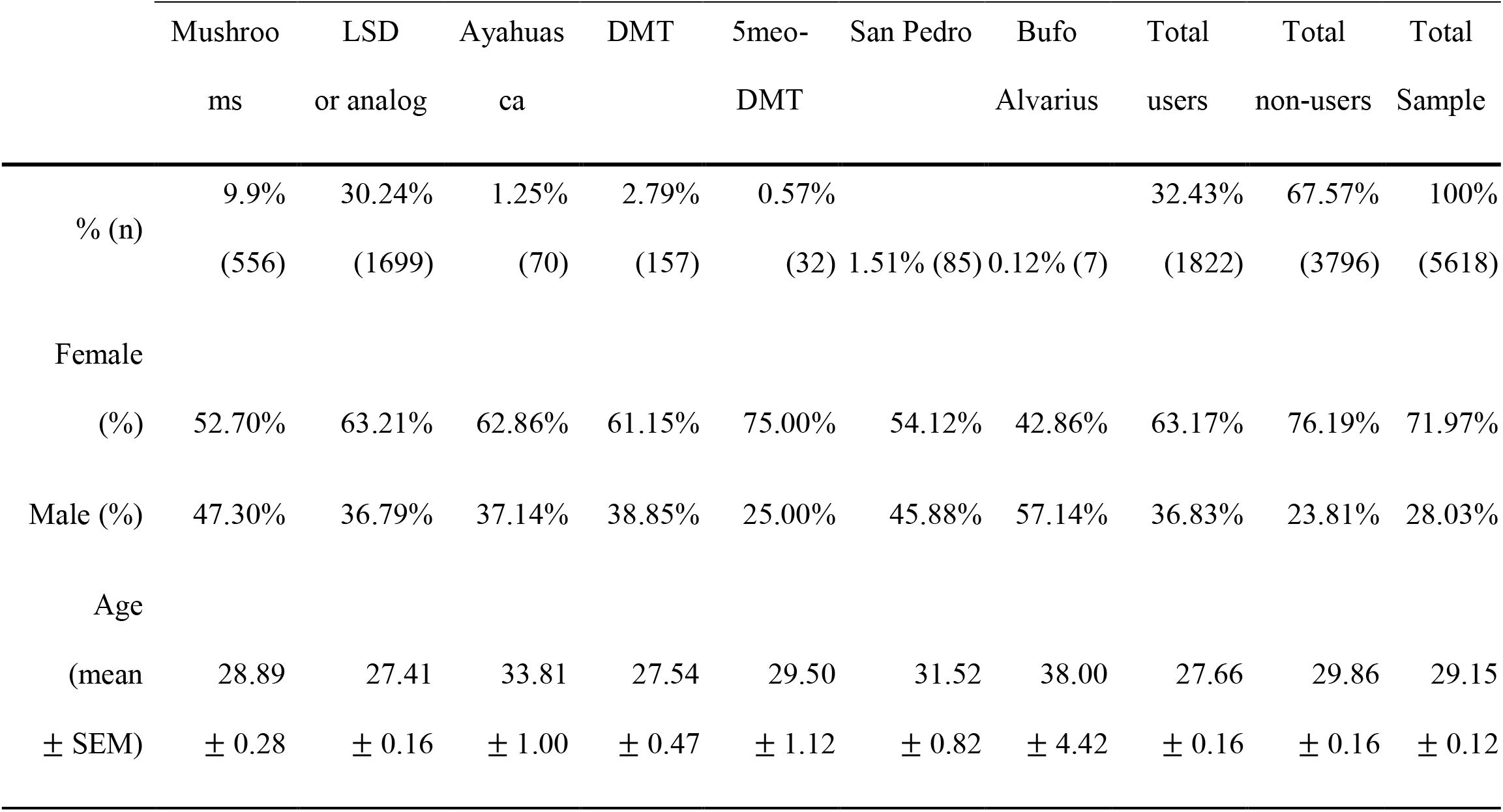
Demographic information corresponding to the subset of 5618 participants who met the inclusion criteria (i.e. Argentinian residents older than 18 years old, with male or female gender identity).

**Table 2.**
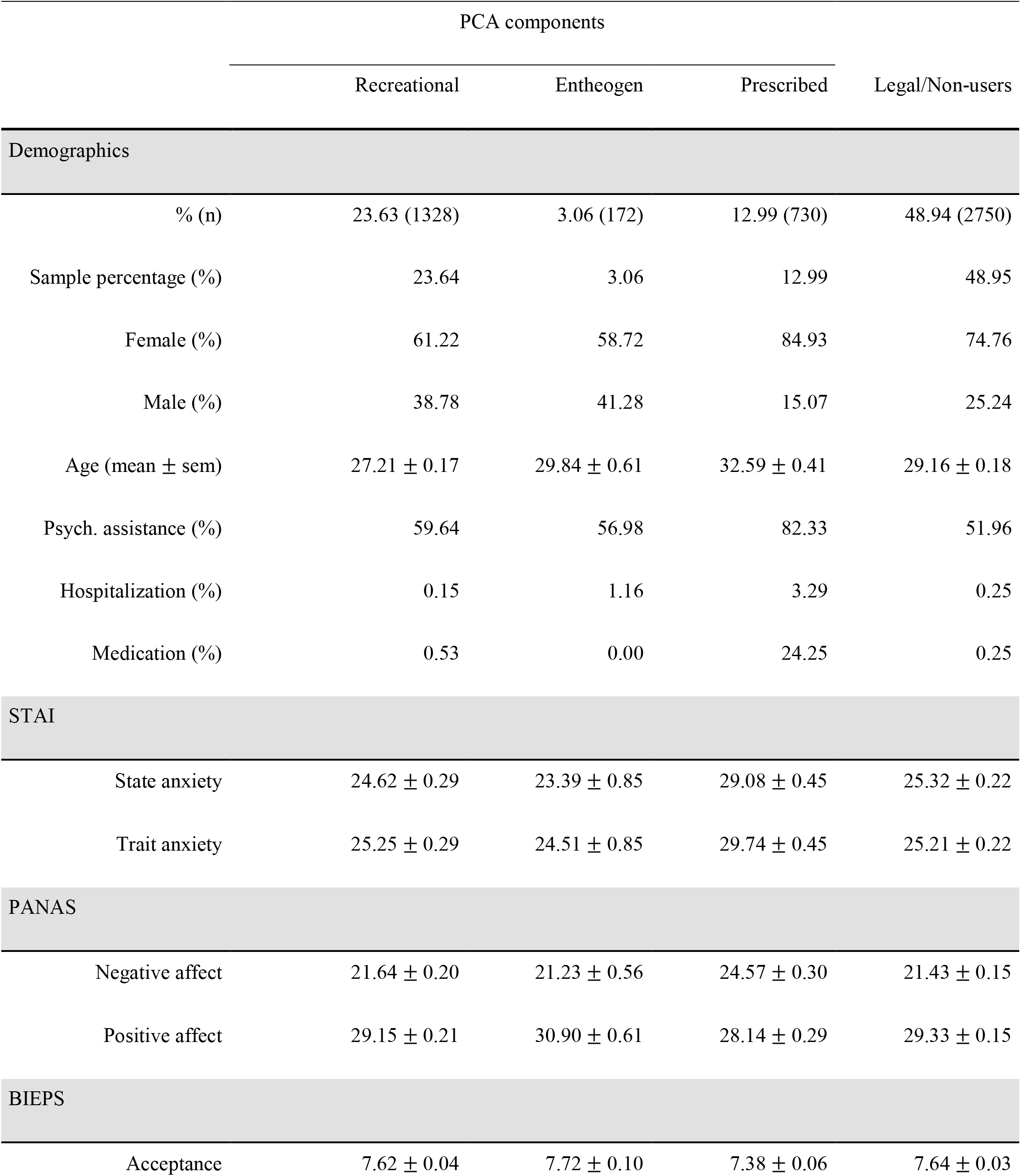
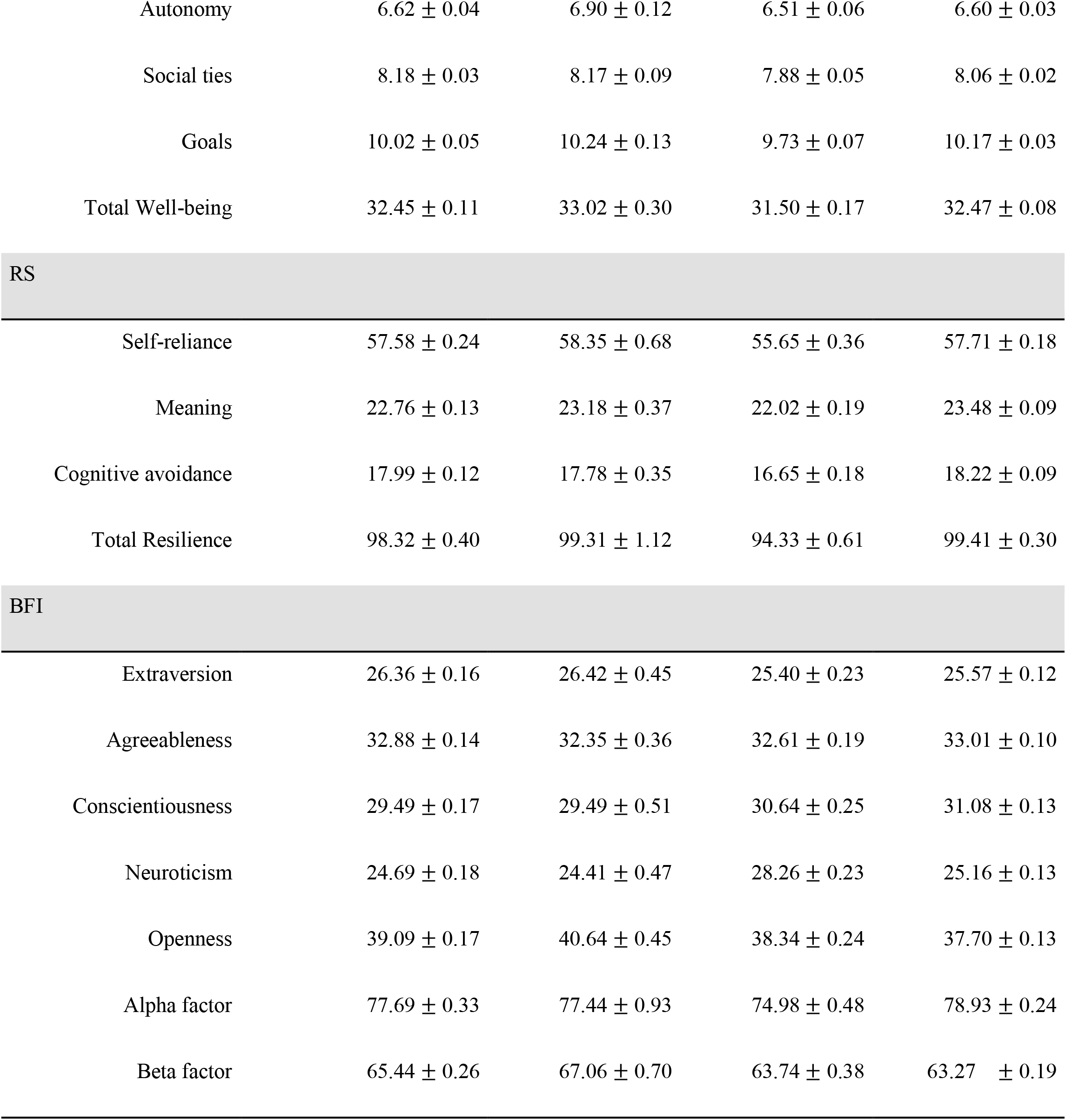
Survey results grouped by principal component scores. Psychometric scales and subscales results (STAI, PANAS, BIEPS, RS, BFI) are presented as mean ± standard error.

Figure 1 summarizes the results of the BFI questionnaire (panel A), and of the STAI, PANAS, BIEPS and RS scales (panel B). The latter were first transformed to Z-scores, so that scores below/above zero indicate values that are below/above the regional averages. We observed that positive affect (PA), well-being (W) and resilience (R) scores were considerably below zero, which we can interpret as a potential effect of the COVID-19 pandemic and its consequences. Figures 1D and 1E show the same information as in panels A and B, but segmented according to the reported number of psychedelic drug uses. The clearest trends can be observed in panel D for the BFI scores, with the openness (O) and extraversion (E) traits increasing as a function of reported uses, and conscientiousness (C) showing the opposite behavior. Figure 1C presents all pairwise correlations between subscales with |*R*|>0.3. The openness and agreeableness dimensions of the BFI questionnaire were not correlated with any of the other scales, while expected positive correlations between state/trait anxiety, negative affect and neuroticisim were observed. Resilience and well-being were positively correlated (and negatively correlated with state/trait anxiety, negative affect and neuroticism), and also presented positive correlations with positive affect, extroversion, and conscientiousness.

**Figure 1.**
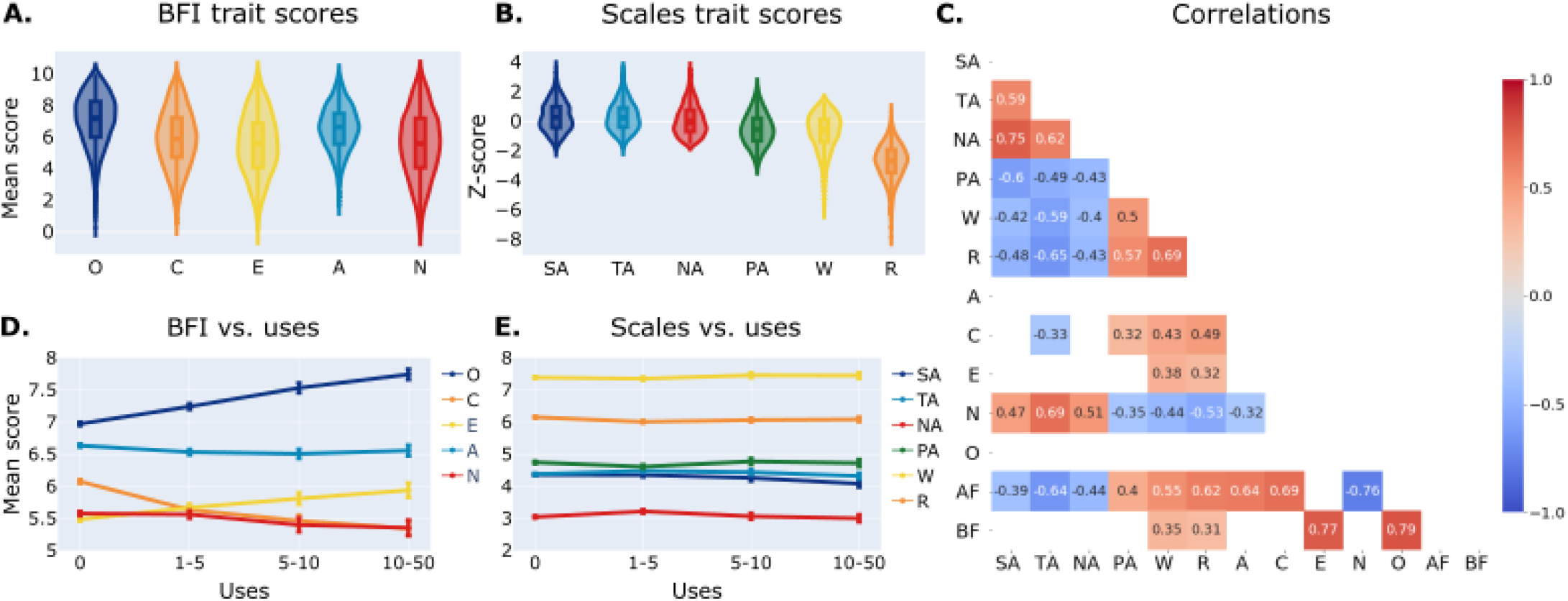
Summary of the BFI questionnaire (panel A), the STAI, PANAS, BIEPS and RS scales (panel B), both as a function of the reported number of psychedelic drug uses (panels D and E), and all pairwise correlations between subscales with |*R*|>0.3. Abbreviations: O (Openness), C (Conscientiousness), E (Extraversion), A (Agreeability), N (Neuroticism), SA (State anxiety), TA (Trait anxiety), NA (Negative affect), PA (Positive affect), W (Well-being), R (Resilience), AF (Alpha factor), BF (Beta factor).

We then performed an exploratory analysis by grouping all scores from users who reported experience with each drug and comparing those scores with the rest of the sample using Student’s T tests. Results are shown in Fig. 2. A double dissociation effect is apparent: some psychedelic drugs (mainly psilocybin mushrooms but also DMT and ayahuasca) were associated with lower scores of dimensions linked to mental health impairment (state/trait anxiety, negative affect) and with higher scores of dimensions linked to well-being and resilience (mainly positive affect, autonomy, social ties and well-being); conversely, other drugs (including prescription drugs, caffeine, alcohol, tobacco and cannabis) were linked to higher scores of dimensions linked to mental health impairment, and lower scores of dimensions linked to well-being and resilience. The exception was MDMA, which was associated with higher scores of the BIEPS social ties dimension.

**Figure 2.**
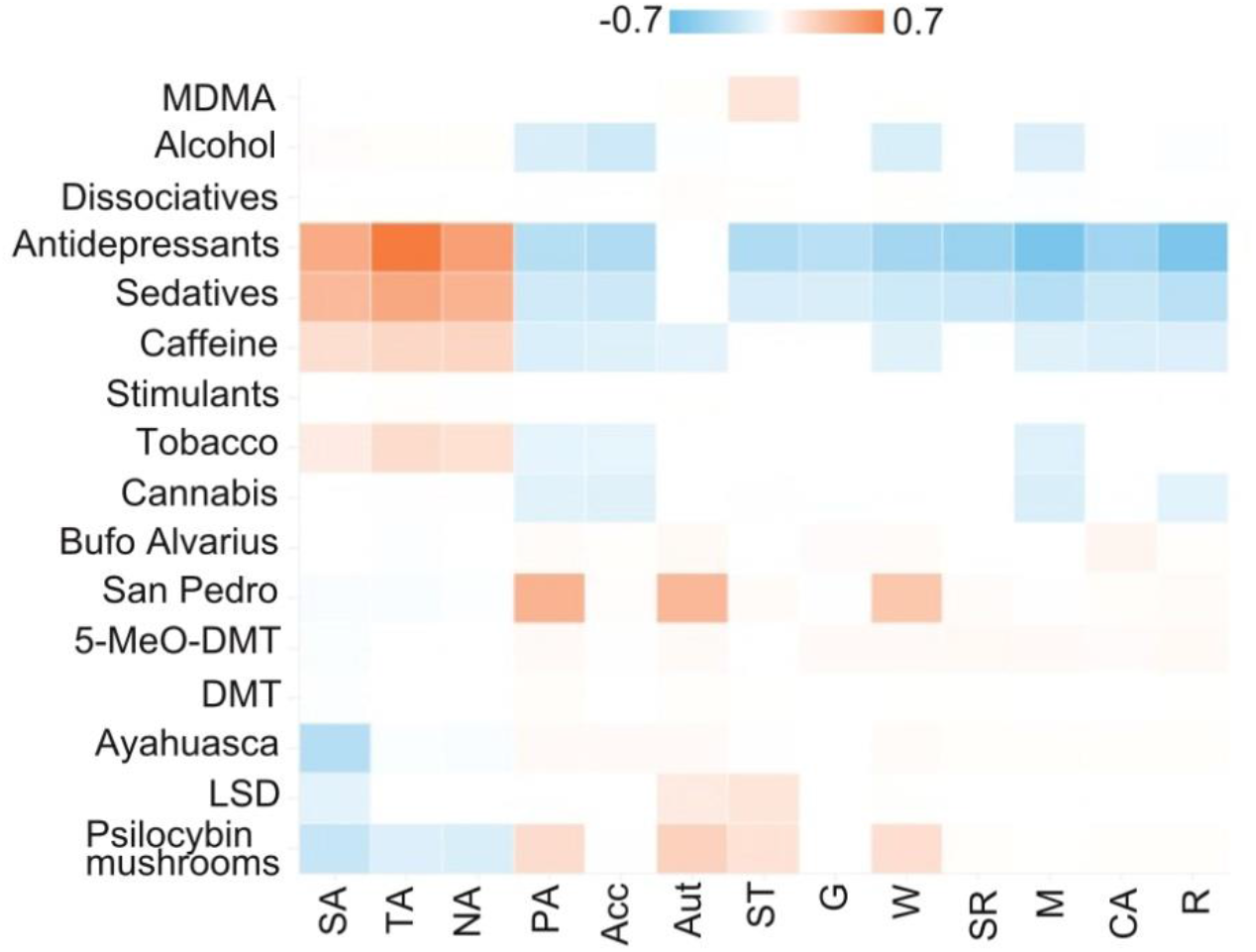
Exploratory analysis comparing questionnaire scores corresponding to individual drugs vs. all others. Each entry in the matrix corresponds to the effect size (Cohen’s d) for the comparison of the score (columns) for the drug (rows) vs. all other drugs grouped together. Only significant entries are shown (p<0.05, Bonferroni corrected for multiple comparisons). Abbreviations: SA (State anxiety), TA (Trait anxiety), NA (Negative affect), PA (Positive affect), Acc (Acceptance), Aut (Autonomy), ST (Social ties), G (Goals), W (Well-being), SR (Self-reliance), M (Meaning), CA (Cognitive avoidance), R (Resilience).

We note that this analysis is exploratory since it does not take into account the possibility of a single participant declaring past experience with multiple drugs. To overcome this limitation, we applied PCA to divide the sample into groups of subjects with different drug use profiles. Each group was created based on the scores along three principal components: LSD, MDMA, psilocybin mushrooms and dissociative drugs (“recreational”, i.e. drugs that are frequently consumed in a recreational setting), DMT, ayahuasca, San Pedro (*Echinopsis pachanoi*), bufo alvarius (*Incilius alvarius*) and 5-MeO-DMT (“entheogen”, i.e. psychedelic compounds that are frequently consumed in religious or ceremonial settings), and antidepressants, antipsychotics and sedatives (“prescription”). The resulting components are shown in Fig. 3A. To ensure that these groups were mutually exclusive, participants with high scores in the “entheogen” and “recreational” components were only included in the latter, e.g. a user with extensive lifetime use of LSD and ayahuasca would only be included in the “entheogen” group. This reflects the observation that most users with lifetime use of drugs in the “entheogen” component also reported experiences with drugs in the “recreational” component, but not vice versa. Subjects presenting high scores for drugs in the “prescription” component were included in the corresponding group, regardless of their scores in the other two components. Finally, we created a group for participants who only declared past experiences with legal drugs, such as caffeine, alcohol and tobacco (“legal/non-users”). 11.36% of the sample did not clearly fit in any group based on the PCA scores, and hence was discarded from subsequent analysis.

**Figure 3.**
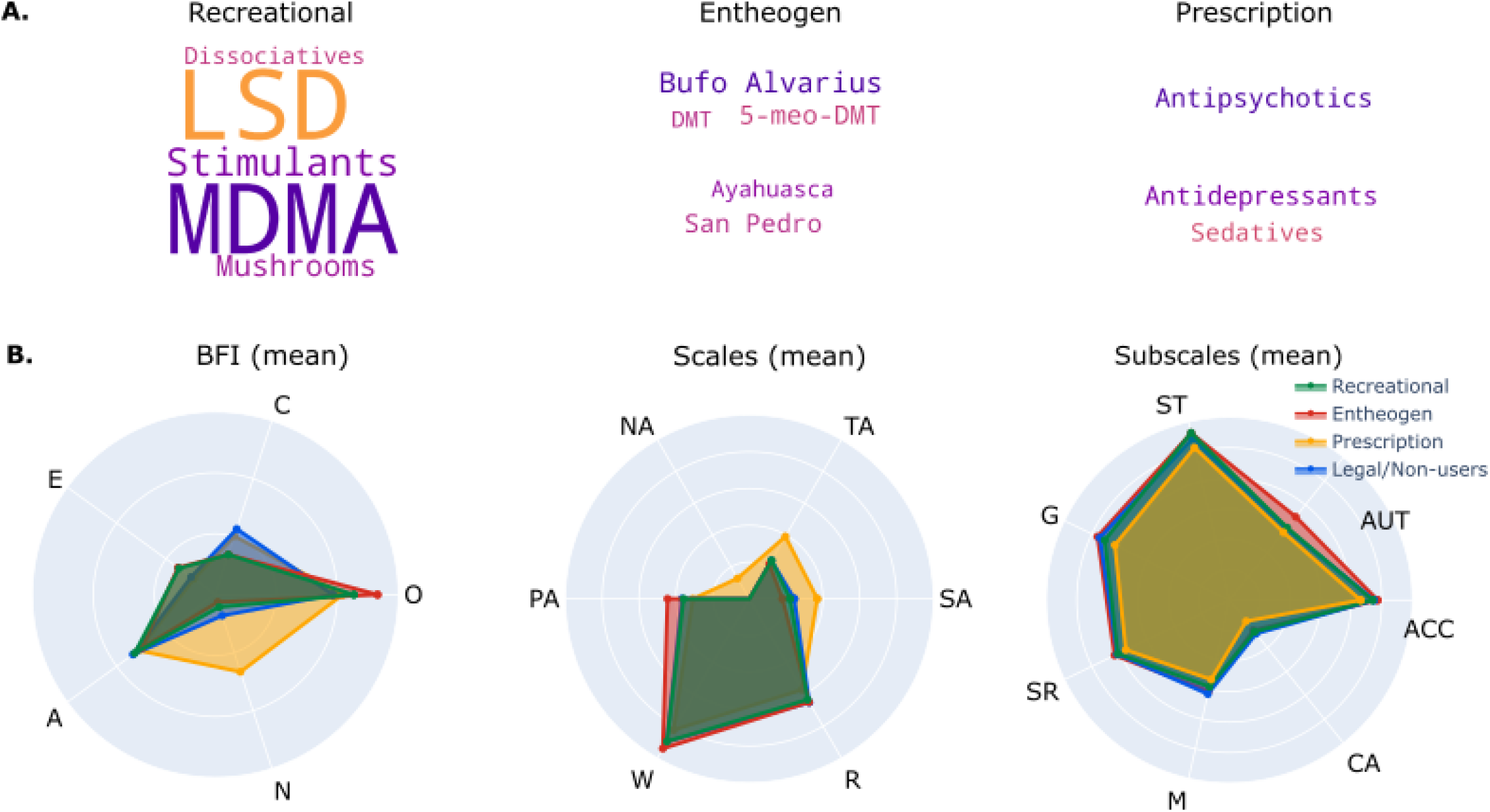
Principal components. A) Word clouds representing the drugs included in the different groups, with word size indicative of the weight of the drug in the corresponding principal component. B) Radar plots showing BFI scores (left), psychometric scale scores (middle) and their subscales (right) for participants belonging to each group, plus a group of participants who only declared past use of legal drugs (e.g. caffeine, alcohol, tobacco). Abbreviations: O (Openness), C (Conscientiousness), E (Extraversion), A (Agreeability), N (Neuroticism), SA (State anxiety), TA (Trait anxiety), NA (Negative affect), PA (Positive affect), W (Well-being), R (Resilience), ACC (Acceptance), AUT (Autonomy), ST (Social ties), G (Goals), SR (Self-reliance), M (Meaning), CA (Cognitive avoidance).

We first applied an ANOVA test to find a significant effect (p<0.05, Bonferroni corrected) of user group in all BFI scores and psychometric scales except for the agreeableness personality trait (p = 0.121).

We applied pairwise Student’s t-test to determine whether the variables plotted in Fig. 3B presented significant differences between the four groups of participants. The results of this analysis are shown in Fig. 4 (state/trait anxiety, BFI, PANAS, BIEPS and RS) and Fig. 5 (alpha and beta factors). Figure 5C also presents the second order personality factors alpha and beta as a function of the reported number of lifetime psychedelic experiences.

**Figure 4.**
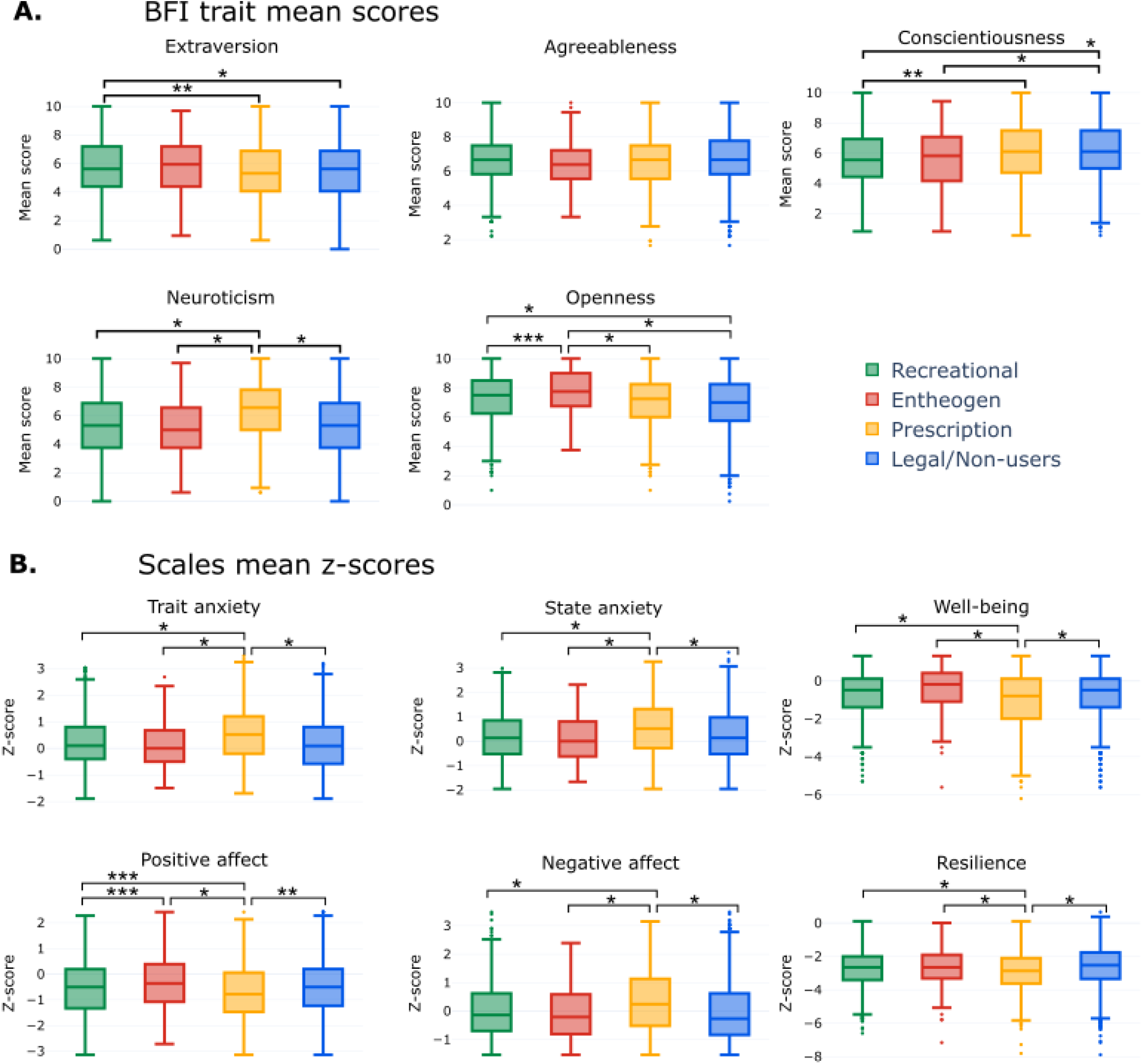
Comparison of psychometric scales and subscales between groups of participants. A) Comparison of BFI dimensions. B) Comparison of trait/state anxiety, positive and negative affect, well-being and resilience. *p<0.05 (Bonferroni corrected), **p<0.01, ***p<0.05.

**Figure 5.**
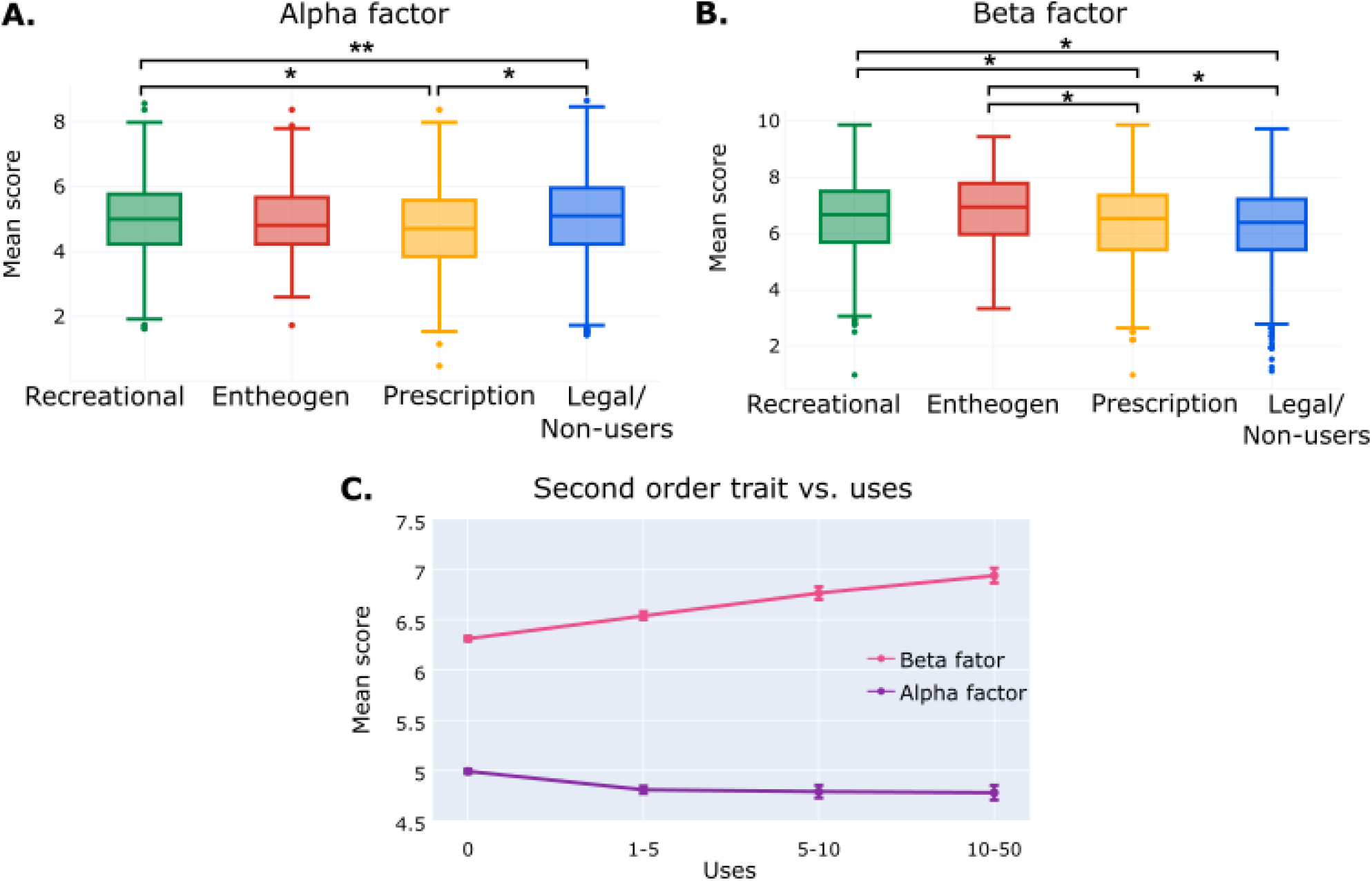
Comparison of second order personality traits between groups of participants. A) Comparison of alpha factor scores. B) Comparison of beta factor scores. C) Alpha and beta factor vs. reported number of lifetime psychedelic experiences. *p<0.05 (Bonferroni corrected), **p<0.01, ***p<0.05.

The “recreational” group presented higher extraversion and lower conscientiousness scores than the group of non-users, while both “recreational” and “entheogens” resulted in increased openness scores, as expected from previous studies. Also as expected, neuroticism was increased in the “prescription” group relative to all others. No differences were found for the agreeableness personality dimension. Subjects in the “entheogen” group presented higher openness than those in the “recreational” group (Fig. 4A). All measures positively associated with mental health were significantly lower in the “prescription” group; also, we observed higher positive affect in the “entheogen” group compared to all others (Fig. 4B). Finally, the alpha factor was lower for the “prescription” group compared to all others (Fig. 5A), and the beta factor was higher for the “recreational” and “entheogen” groups compared to the other two groups (Fig. 5B). Also, the beta factor increased with the reported number of lifetime psychedelic experiences (Fig. 5C).

## Discussion

The current study investigated the relationship between lifetime use of psychoactive drugs and reported levels of anxiety, positive and negative affect, well-being, resilience, and personality traits. We focused our analysis on the possibility that psychedelic drugs could confer sustained benefits, which are manifested as better mental health indicators during the ongoing COVID-19 pandemic.

Concerning the general mental health status of the sample, after scoring positive and negative scales and transforming them to z-scores we observed reduced values, particularly in resilience (−2.74), but also in well-being (−0.74) and PANAS positive affect (−0.54), and higher values in STAI state anxiety (0.3), STAI trait anxiety (0.28), and PANAS negative affect (0.06) (Fig 4, panel B). We expected an effect on mental health indicators as a consequence of the social isolation and uncertainty associated with the COVID-19 outbreak. This result is in line with preliminary reports showing serious consequences in the mental health of the general population during the pandemic (Pfefferbaum and North, 2020; Zacher and Rudolph, 2020; Gallagher et al., 2020, Brooks et al., 2020).

We observe that our results successfully replicated known results concerning the relationship between personality traits and different mental health indicators (Widiger and Trull, 1992; Widiger and Costa, 1994, Mineka, Watson and Clark, 1998; Kotov et al., 2010; Beck, Davis and Freeman, 2015). Figure 1C confirms the positive correlation between BIEPS, RS, and PANAS positive affect scores, and negative correlations between STAI trait and state, and PANAS negative affect (Hu et al., 2015). Concerning personality traits, neuroticism correlated both with temporary psychological states (i.e. negative affect, state anxiety) and more stable personality characteristics (i.e. trait anxiety), confirming a significant association between this trait and negative emotion (Costa and McCrae, 1977; Costa and McCrae, 1980; John et al., 2008). Higher order traits also behaved as expected, beta factor being correlated with the well-being and resilience scores, and alpha factor behaving opposite to the neuroticism trait, i. e. exhibiting similar correlations of the opposite sign (Bäckström et al., 2007; Kardum and Hudek-Knezevic, 2012).

We investigated the relationship between self-reported drug use and the psychometric questionnaire scores by first applying principal component analysis to group subjects according to their experiences with psychoactive drugs.. These groups reflected how the use of drugs was clustered in our sample: the first component included mainstream psychedelic, entactogen and dissociative drugs, most of which are consumed in recreational contexts; the second group only included serotonergic psychedelics that are commonly consumed in religious or ceremonial context; the final group included prescription psychoactive drugs such as antidepressants, antipsychotics and sedatives. This classification was useful to highlight the specific effects of certain psychedelic drugs, which likely transcend their pharmacological action and emerge as a consequence of interactions with contextual factors (i.e. set and setting) (Kazdin, 2007, Mulder et al 2017, Carhart-Harris et al., 2018).

We confirmed that experience with psychedelic drugs was associated with changes in personality traits indexing the experience of novelty (MacLean et al., 2011; Lebedev et al., 2016; Bouso et al., 2018; Erritzoe et al., 2018; Erritzoe et al., 2019). Openness refers to active curiosity in the intellectual domain, while extraversion, represents openness behavioral counterpart but oriented to the material world, involving an active type of curiosity that includes (but is not limited to) social interactions (DeYoung et al., 2002). Both openness and extraversion are functional characteristics, since a person who faces the environment in a positive way is more likely to obtain reinforcement from the interaction and thus adapt his or her behavior accordingly, suggesting a link between these traits and serotonergic and dopaminergic circuits (Ashby and Isen, 1999; Depue and Collins, 1999; DeYoung et al., 2002). As shown in Fig. 1D, the reported number of psychedelic drug uses was positively correlated with openness and extraversion; furthermore, lifetime use of psychedelic were linked to significant increases in these traits (with the exception of the extraversion trait for the “entheogen” group). Although these results are particularly interesting, due to the nature of our study we cannot confirm whether this increment was caused by lifetime use of psychedelic drugs, or due to different pre-existing personality traits psychedelic drug users.

Concerning second order personality traits, alpha and beta factors presented significant increases in the groups of psychedelic users, and the reported number of psychedelic uses correlated with both traits. The alpha factor is obtained as a combination of agreeableness, conscientiousness and the inverse of neuroticism, and has been interpreted as a measure of social desirable traits (Bäckström et al., 2007; Kardum and Hudek-Knezevic, 2012). This factor has significantly increased in the “recreational” group but not in the “entheogen” group. The beta factor is obtained as a combination of extraversion and openness and has been interpreted as a striving for self-assertion and self-expansion (Bäckström et al., 2007; Kardum and Hudek-Knezevic, 2012). This factor was significantly increased in both the “recreational” and “entheogen” group, and also correlated positively with the reported number of psychedelic drug uses. Alternatively, the alpha and beta factors have been interpreted in terms of “stability” and “plasticity”, respectively (Digman, 1997; DeYoung et al., 2002). The positive association between lifetime use of psychedelics and these personality traits suggests enhanced resilience and well-being in the light of challenging situations (Figure 1C).

Certain considerations can be drawn in regards to the link between psychedelics and changes in personality traits. First, since some of these changes could be related to 5-HT_2A_ receptor expression and activation (Kalbitzer et al., 2009), it is hypothesized that a pharmacological interaction occurring in a sustained way could modify genetic expression and thus promote stable modifications in the personality of the users and therefore also in their behavioral patterns (Bouso et al., 2018). In this case, a causal link could exist between the changes in personality trait and the reported number of psychedelic drug uses (Figure 5C). Second, this could also offer psychotherapeutic potential by external modulation of personality traits. This can be considered both an end by itself (e.g. in the case of certain disorders), or an intermediate objective if the therapeutic goal is to make the patient more flexible to work on psychopathological aspects underlying their personality style (e.g., mood disorders like depression, caused by personality disorders like avoidant personality disorder).

Additionally, all the personality traits evaluated in this study (i.e., extraversion, agreableness, conscientiousness, neuroticism, openness), and their second order factors (i.e., alpha and beta) are psychological representations whose utility is to model the most stable and durable cognitive style of individuals. As a stable factor, personality bears a large part of the variance of the temporal states of mind (e.g., the clinical association between psychopathological conditions and the neuroticism trait). Likewise, these temporary states are caused by the cognitive evaluation resulting from the interaction with the environment, and influence secondary constructions that involve self-evaluation (i.e., well-being and resilience). It is therefore appropriate to propose a logical order in which the variables of this study are linked: the most stable factors (personality traits and anxiety as a trait) influence the cognitive style that is responsible for interpreting and processing reality. In turn, what is interpreted affects emotions, which can be considered as temporary cognitive states (anxiety, positive affect, negative affect). The sustained summation of these states makes up, among other things, the self-perceived processes of well-being and resilience.

We must clarify certain limitations arising from methodology. First, it is not possible to corroborate the information given by the participants, especially concerning the identity and dose of the psychoactive drugs they consumed. Second, it is possible that variables outside the scope of our survey are influencing the results we obtained. Concerning this limitation, our principal component analysis aimed to alleviate the effect of confounds caused by poly-drug use by dividing the sample into disjoint groups depending on their principal component values. Finally, it was not possible to draw causal inferences due to our study design.

In summary, we performed a survey to investigate the relationship between mental health, personality and past drug use during the COVID-19 pandemic. Our results do not suggest an association between past psychedelic use and impaired mental health indicators; on the contrary, we found evidence supporting a more resilient and stable personality structure in those subjects who reported repeated use of certain psychedelic compounds. This study adds to the existing literature on the relationship between mental health and lifetime psychedelic use by investigating self-reported measures of well-being during a highly challenging situation known to cause adverse psychological responses. Future studies should investigate with more detail whether our results can be attributed to the long-term changes induced by psychedelics, and how these changes relate to the available evidence concerning the potential use of psychedelics in the treatment of psychiatric disorders.

## Data Availability

Data not available yet.

## Acknowledgments

This study has been funded by grant COVID-19 IP 264 by Agencia Nacional de Promoción de la Investigación, el Desarrollo Tecnológico y la Innovación.

