## Supplementary material for "Mental health, personality and lifetime psychedelic use during the COVID-19 pandemic"

**Table S1.** Survey results grouped by drugs.

| | Mushrooms | LSD or analog | Ayahuasca | DMT | 5meo-DMT | San Pedro | Bufo Alvarius | Total users | Total non-users | Total sample |
|---|---|---|---|---|---|---|---|---|---|---|
| Demographics | | | | | | | | | | |
| % (n) | 9.9 (556) | 30.24 (1699) | 1.25 (70) | 2.79 (157) | 0.57 (32) | 1.51 (85) | 0.12 (7) | 32.43 (1822) | 67.57 (3796) | (5618) |
| Female (%) | 52.70 | 63.21 | 62.86 | 61.15 | 75.00 | 54.12 | 42.86 | 63.17 | 76.19 | 71.97 |
| Male (%) | 47.30 | 36.79 | 37.14 | 38.85 | 25.00 | 45.88 | 57.14 | 36.83 | 23.81 | 28.03 |
| Age (mean ± sem) | 28.89 ±0.28 | 27.41 ± 0.16 | 33.81 ± 1.00 | 27.54 ± 0.47 | 29.50 ± 1.12 | 31.52 ± 0.82 | 38.00 ± 4.42 | 27.66 ± 0.16 | 29.86 ± 0.16 | 29.15 ± 0.12 |
| Psych. assist. (%) | 66.37 | 66.22 | 68.57 | 66.24 | 68.75 | 62.35 | 71.43 | 65.81 | 59.19 | 61.34 |
| Hospitalization (%) | 1.26 | 1.53 | 2.86 | 3.18 | 9.38 | 1.18 | 14.29 | 1.54 | 1.03 | 1.19 |
| Medication (%) | 5.76 | 5.59 | 10.00 | 6.37 | 9.38 | 2.35 | 28.57 | 5.43 | 5.64 | 5.57 |
| STAI | | | | | | | | | | |
| State anxiety | 24.74 ±0.48 | 25.75 ± 0.27 | 23.97 ± 1.37 | 25.87 ± 0.97 | 25.88 ± 2.22 | 24.06 ± 1.32 | 26.86 ± 3.98 | 25.67 ± 0.26 | 26.17 ± 0.19 | 26.01 ± 0.15 |
| Trait anxiety | 25.59 ± 0.48 | 26.78 ± 0.27 | 24.86 ± 1.37 | 27.03 ± 0.97 | 27.66 ± 2.22 | 24.54 ± 1.32 | 28.14 ± 3.98 | 26.61 ± 0.26 | 26.26 ± 0.19 | 26.37 ± 0.15 |

| | | | | | | | | | | |
|---|---|---|---|---|---|---|---|---|---|---|
| PANAS | | | | | | | | | | |
| Negative affect | 21.81 ± 0.32 | 22.66 ± 0.19 | 21.46 ± 0.96 | 22.49 ± 0.59 | 24.09 ± 1.35 | 21.93 ± 0.88 | 22.14 ± 3.36 | 22.57 ± 0.18 | 22.18 ± 0.13 | 22.31 ± 0.10 |
| Positive affect | 29.35 ± 0.33 | 28.63 ± 0.19 | 31.07 ± 0.96 | 28.97 ± 0.66 | 29.34 ± 1.61 | 31.02 ± 0.89 | 30.14 ± 3.83 | 28.71 ± 0.18 | 28.99 ± 0.13 | 28.90 ± 0.11 |
| BIEPS | | | | | | | | | | |
| Acceptance | 7.59 ± 0.06 | 7.50 ± 0.03 | 7.69 ± 0.16 | 7.50 ± 0.12 | 7.44 ± 0.25 | 7.72 ± 0.13 | 8.14 ± 0.26 | 7.52 ± 0.03 | 7.56 ± 0.02 | 7.55 ± 0.02 |
| Autonomy | 6.77 ± 0.07 | 6.61 ± 0.04 | 7.09 ± 0.19 | 6.84 ± 0.13 | 6.91 ± 0.27 | 6.99 ± 0.16 | 7.43 ± 0.53 | 6.62 ± 0.04 | 6.58 ± 0.03 | 6.59 ± 0.02 |
| Social ties | 8.18 ± 0.05 | 8.12 ± 0.03 | 8.41 ± 0.11 | 7.94 ± 0.11 | 7.69 ± 0.27 | 8.32 ± 0.11 | 7.86 ± 0.55 | 8.12 ± 0.03 | 8.02 ± 0.02 | 8.05 ± 0.02 |
| Goals | 9.92 ± 0.08 | 9.95 ± 0.04 | 10.17 ± 0.23 | 10.02 ± 0.14 | 10.41 ± 0.28 | 10.26 ± 0.19 | 10.71 ± 0.36 | 9.97 ± 0.04 | 10.05 ± 0.03 | 10.03 ± 0.02 |
| Total Well-being | 32.46 ± 0.17 | 32.19 ± 0.10 | 33.36 ± 0.44 | 32.29 ± 0.34 | 32.44 ± 0.75 | 33.28 ± 0.40 | 34.14 ± 1.28 | 32.23 ± 0.09 | 32.21 ± 0.07 | 32.22 ± 0.05 |
| RS | | | | | | | | | | |
| Self-reliance | 57.55 ± 0.38 | 56.81 ± 0.22 | 58.31 ± 1.01 | 56.99 ± 0.77 | 57.06 ± 1.76 | 57.92 ± 0.90 | 60.43 ± 3.75 | 56.89 ± 0.21 | 57.16 ± 0.16 | 57.07 ± 0.13 |
| Meaning | 22.57 ± 0.22 | 22.21 ± 0.12 | 23.10 ± 0.59 | 22.48 ± 0.39 | 23.56 ± 0.86 | 22.72 ± 0.52 | 22.71 ± 2.36 | 22.27 ± 0.12 | 23.09 ± 0.08 | 22.82 ± 0.07 |

| | | | | | | | | | | |
|---|---|---|---|---|---|---|---|---|---|---|
| Cognitive avoidance | 17.55 ± 0.20 | 17.44 ± 0.11 | 17.61 ± 0.59 | 17.28 ± 0.40 | 17.31 ± 0.89 | 17.48 ± 0.46 | 20.43 ± 1.07 | 17.52 ± 0.11 | 17.88 ± 0.08 | 17.76 ± 0.06 |
| Total Resilience | 97.67 ± 0.66 | 96.4 7 ± 0.37 | 99.03 ± 1.79 | 96.75 ± 1.23 | 97.94 ± 2.90 | 98.12 ± 1.45 | 103.57 ± 6.16 | 96.68 ± 0.36 | 98.12 ± 0.26 | 97.66 ± 0.21 |
| BFI | | | | | | | | | | |
| Extraversion | 26.59 ± 0.26 | 26.41 ± 0.15 | 26.93 ± 0.65 | 26.81 ± 0.47 | 26.31 ± 0.97 | 26.60 ± 0.64 | 26.71 ± 2.24 | 26.39 ± 0.14 | 25.56 ± 0.10 | 25.83 ± 0.08 |
| Agreeableness | 32.54 ± 0.21 | 32.46 ± 0.12 | 32.81 ± 0.61 | 32.00 ± 0.39 | 32.00 ± 0.86 | 33.01 ± 0.48 | 33.14 ± 0.86 | 32.51 ± 0.12 | 32.88 ± 0.08 | 32.76 ± 0.07 |
| Conscientiousness | 29.29 ± 0.28 | 28.88 ± 0.16 | 30.11 ± 0.81 | 28.61 ± 0.52 | 29.81 ± 1.22 | 28.26 ± 0.74 | 26.14 ± 2.79 | 28.96 ± 0.15 | 30.87 ± 0.11 | 30.25 ± 0.09 |
| Neuroticism | 24.97 ± 0.28 | 25.66 ± 0.16 | 24.66 ± 0.73 | 25.70 ± 0.50 | 25.53 ± 1.32 | 24.18 ± 0.65 | 23.71 ± 2.20 | 25.57 ± 0.15 | 25.84 ± 0.11 | 25.75 ± 0.09 |
| Openness | 40.30 ± 0.25 | 39.60 ± 0.15 | 41.24 ± 0.71 | 41.34 ± 0.44 | 41.81 ± 0.83 | 41.62 ± 0.59 | 42.00 ± 2.30 | 39.60 ± 0.14 | 37.89 ± 0.11 | 38.44 ± 0.09 |
| Alpha factor | 76.87 ± 0.54 | 75.68 ± 0.31 | 78.27 ± 1.40 | 74.91 ± 1.00 | 76.28 ± 2.19 | 77.09 ± 1.24 | 75.57 ± 3.88 | 75.90 ± 0.30 | 77.91 ± 0.21 | 77.26 ± 0.17 |
| Beta factor | 66.89 ± 0.39 | 66.00 ± 0.23 | 68.17 ± 1.07 | 68.15 ± 0.69 | 68.12 ± 1.42 | 68.22 ± 0.95 | 68.71 ± 3.77 | 65.99 ± 0.22 | 63.45 ± 0.16 | 64.27 ± 0.13 |

**Table S1**. The information shown corresponds to the sample subset (participants older than 18 years old, Argentinians, who live in Argentina, with male or female gender identity). Consumers in each user group are not mutually exclusive with the other groups (users in each group may have used more than one drug).
